# Machine Learning Applied to Routine Blood Tests and Clinical Metadata to Identify and Classify Heart failure

**DOI:** 10.1101/2021.07.26.21261115

**Authors:** Nick James, Lianna Gerrish, Nikita Rokotyan, Patrick A. Gladding

## Abstract

**Introduction:** We applied machine learning (ML) to routine bloods, then to advanced haematology data from a full blood count (rawFBC) plus biochemistry, to build predictive models for heart failure, which were then used at population scale.

**Methods:** Routine blood results from 8,031 patients with heart failure, with equal number of controls, were used in ML training and testing datasets (Split 80:20). NT-proBNP was used for diagnostic comparison. rawFBC metadata was used in a dataset of 698 patients, 314 of whom had heart failure, to train and test ML models (Split 70:30) from rawFBC, rawFBC plus biochemistry and routine bloods. The rawFBC model was used to predict heart failure in a validation dataset of 69,492 FBCs (2.3% heart failure prevalence).

**Results:** Heart failure was predicted from rawFBC and biochemistry versus rawFBC AUROC 0.93 versus 0.91, 95% CI -0.023 to 0.048, P = 0.5, and predicted from routine bloods and NT-proBNP, AUROC 0.87 versus 0.81, 95% CI 0.004 to 0.097, P = 0.03. In the validation cohort heart failure was predicted from rawFBC with AUROC 0.83, 95% CI 0.83 to 0.84, P < 0.001, sensitivity 75%, specificity 76%, PPV 7%, NPV 99.2% (Figure 2). Elevated NT-proBNP (≥ 34 pmol/L) was predicted from rawFBC with AUROC 0.97, 95% CI 0.93 to 0.99, P < 0.0001. Common predictive features included markers of erythropoiesis (red cell distribution width, haemoglobin, haematocrit).

**Conclusion:** Heart failure can be predicted from routine bloods with accuracy equivalent to NT-proBNP. Predictive features included markers of erythropoiesis, with therapeutic monitoring implications.

## Introduction

The term machine learning describes a range of pattern recognition tools which hold considerable promise in the field of health care diagnostics, prognostics and therapeutics. Machine learning has been applied to a number of imaging and diagnostic modalities, including retinal photography (1), electrocardiography (2, 3), and echocardiography (4) to enable cardiovascular disease prediction. Deep learning, a form of machine learning, directed at echocardiography cine images has been shown to be highly accurate at identifying cardiac amyloidosis (4). Integrating both ECG and echocardiography artificial intelligence models provides an even more accurate prediction of the presence of cardiac disease (5, 6). The greatest opportunity for the use of machine learning in healthcare is in its application to low cost, readily available data which has imperceptible patterns hidden within it, which answer unmet clinical needs in healthcare.

In heart failure several machine learning studies have been performed, using data extracted from electronic healthcare records (7-9). Heart failure prognosis has been predicted with a high degree of accuracy using machine learning applied to medical claims data (10). Whilst intriguing, this method is dependent on adequate labelling of ICD10 case data, completeness of clinical records and clinician behaviour, which may not be translatable to other clinical systems or other countries (11). Whilst the potential for this type of bias is considerable with black-box uninterpretable methods such as deep learning, there are several machine learning methods that provide transparency and explainability but with equivalent levels of accuracy.

Although laboratory testing is one of the most ubiquitous, high volume and low cost forms of diagnostic information there is only a recently emerging literature on the use of machine learning applied to lab data (12). One of the most highly evolved tools available, based on machine learning applied to longitudinal full blood count data is the ColonFlag™ Test, designed to flag the presence of colorectal cancer (13). This predictive test has gone from a research discovery to a clinically implemented electronic decision support tool (14). Many studies have demonstrated the value of laboratory data to make not only an accurate prediction of the presence of heart failure but also its prognosis (9, 10, 15).

Unsupervised machine learning, a method applied to unlabelled data to cluster similar patient phenotypes, has been used to identify subtypes of both heart failure to preserved ejection fraction (HFpEF) and heart failure with reduced ejection fraction (HFrEF) (16, 17). Unsupervised machine learning holds the promise of personalising patient care, through the use of targeted therapeutics (18, 19). This work has demonstrated the necessity of not only including a wide breadth of multimodal data, but also standardised inputs, as again this field is suffering from a reproducibility crisis (20). As occurred with genome wide association studies the field of big data in medicine is crying out for a global pooled consortia to ensure machine learning in health care is reproducible and translatable.

We and others have similarly shown the ability for machine learning applied to haematology data to predict the presence of heart failure and biological age (21, 22). The study described here had two parts. Firstly, we set out to integrate haematology and biochemistry data to show the incremental value of adding other sources of laboratory information in the discrimination of patients with heart failure. We then added further clinical metadata, such as conventional ECG parameters and echocardiography measurements, into a more complete holistic picture of patient phenotype. This global metadata was then used to validate previously described HFpEF phenotypes. In the second part we used more granular advanced haematology data to show an ability to identify heart failure at presentation to hospital and how this could be applied across an entire health care system. Lastly, we demonstrate that explainability of machine learning models, a priori, points to potential uses of this method, not only in diagnosis and prognosis, but also therapeutic interventions and monitoring.

## Methods

### Hypothesis

That congestive heart failure (either heart failure with reduced ejection fraction (HFrEF), and heart failure with preserved ejection fraction (HFpEF)) can be predicted from either routine blood investigations e.g. full blood count (FBC) and biochemistry or raw haematology full blood count (rawFBC) data extracted from a haematology analyser/flow cytometer.

### Data collection

This study occurred in two parts. The first part involved the form of a retrospective observational study using data collected as part of routine clinical care from 2008 until 2018 (**E**xtracting Global **Cl**in**i**cal Metadata from Large **P**atient Databa**SE**s (ECLIPSE)). ECLIPSE included extensive patient metadata inclusive of demographics, electrocardiography conventional parameters, laboratory data, inpatient e-prescribing, coronary catheterisation disease coding, CT angiography report extracts, echocardiography measurement data and structured reporting, ICD10 coding, past and future admission and mortality coding. Laboratory data was obtained at a time proximate to the first echocardiogram being performed on a patient with a presumed cardiac diagnosis. This included patients who were both inpatients and in outpatient ambulatory care. Patients with heart failure were identified by any ICD10 coding for heart failure. A matched number of randomly selected patients without heart failure were taken from the ECLIPSE database.

The second part was also a retrospective observational study, (**P**attern **R**ecognition **O**f **G**lobal **R**esponses and **M**onitoring (PROGRAM), though instead this study used advanced haematology raw data as well as biochemistry data. Haematology raw data came from full blood counts (rawFBC) collected between July 1^st^ 2019 and June 8^th^ 2020, some of which occurred during the COVID-19 pandemic. Machine learning (ML) predictions for COVID-19, other infectious diseases and heart failure have been presented previously (22). Apart from COVID-19 these predictive models were developed using the first rawFBC performed during an admission as input data and the primary ICD10 diagnosis as the objective. In this paper we present new ML models for heart failure with the addition of biochemistry data. Comparisons were then possible between each dataset, using each of the predictive models. In addition, we also provide further validation of the same predictive haematology ML model based on an independent dataset of all-comers, undergoing serial FBCs throughout a hospitalisation (Figure 1).

**Figure 1.**
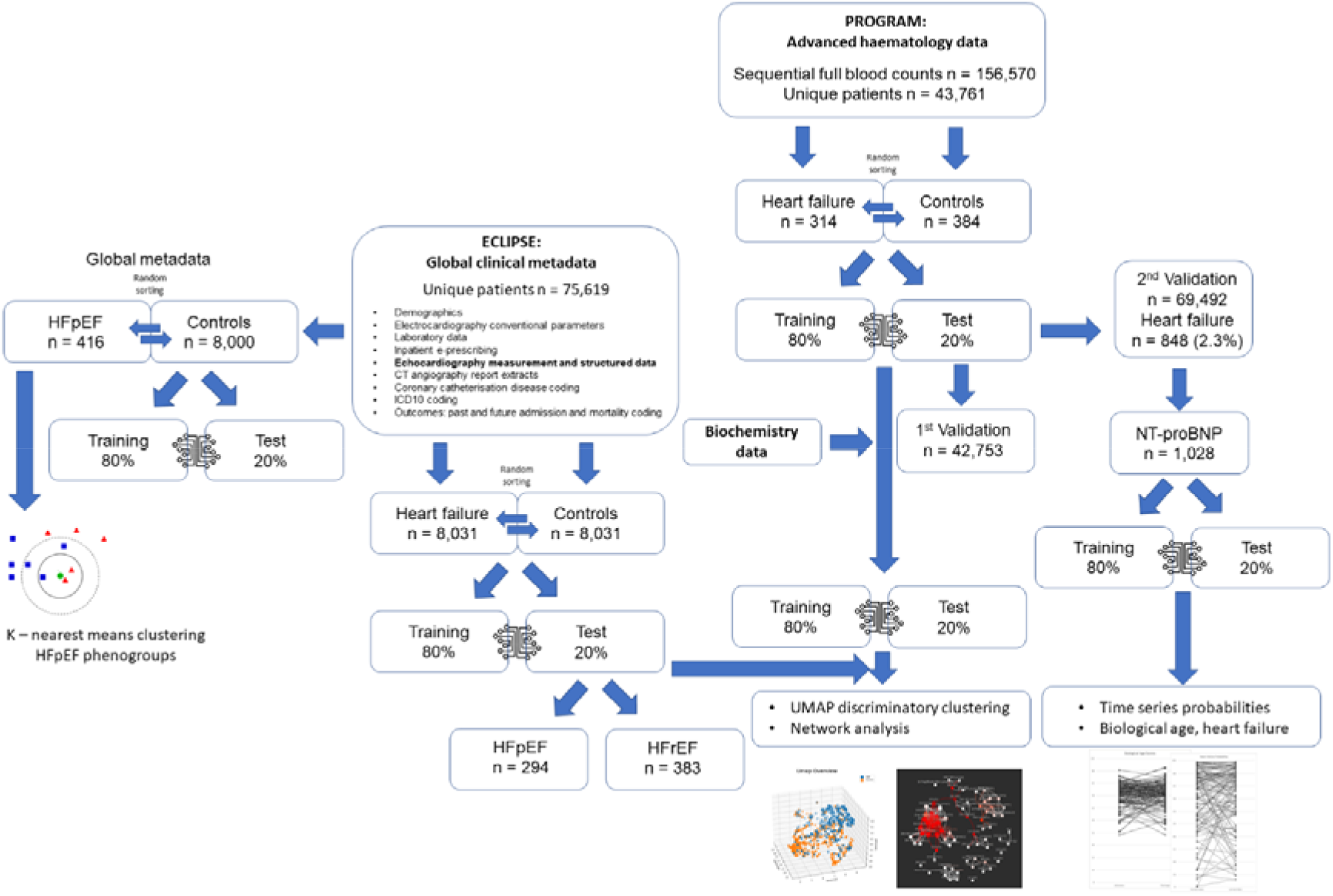
Machine learning application to ECLIPSE and PROGRAM

**Figure 2.**
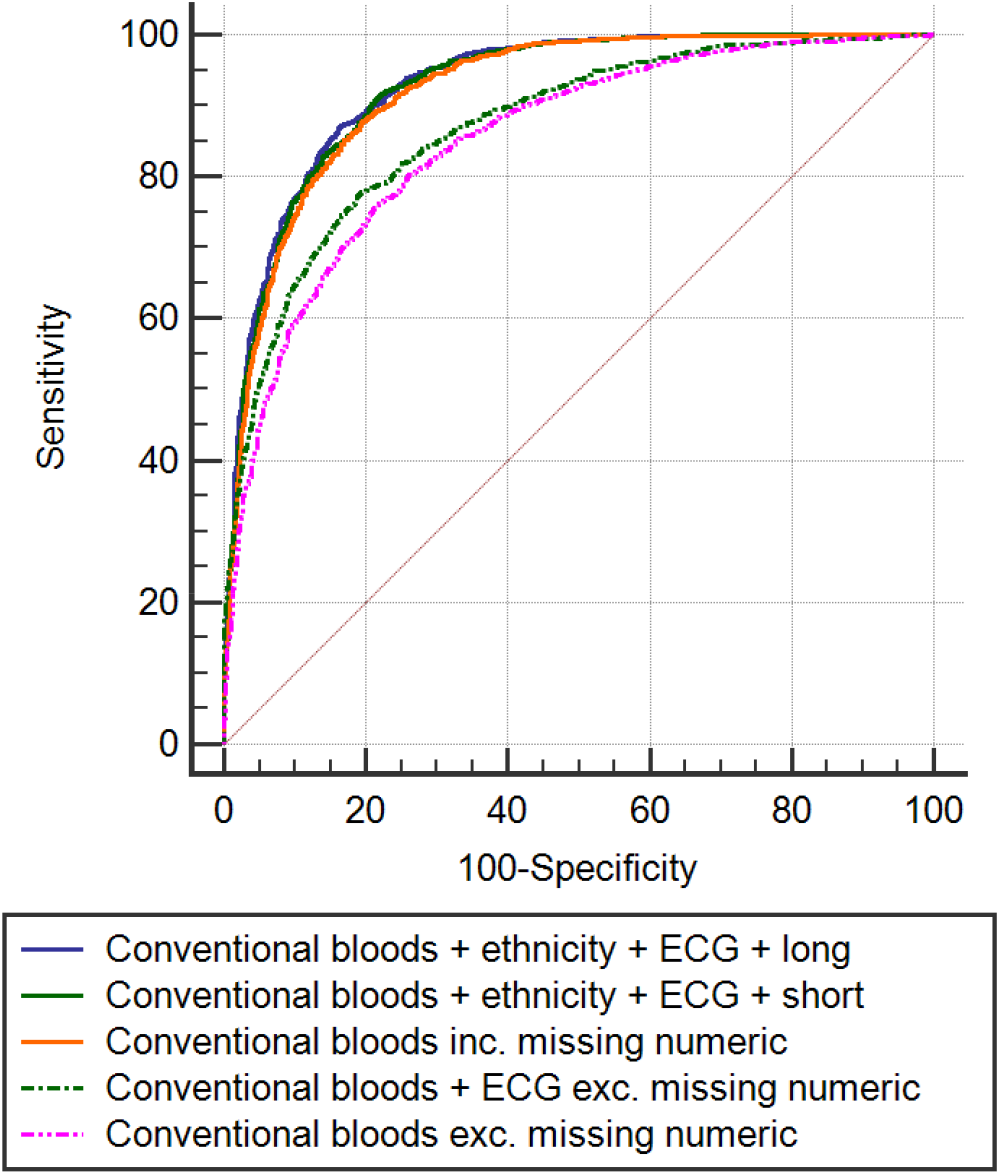
Receiver operator curves comparing ECLIPSE models

Ethics approval was obtained locally from the Research and Knowledge Centre (RM13732 for the ECLIPSE study for standard haematology and biochemistry data), and from the regional HDEC ethics committee (20/CEN/162 for the PROGRAM study for haematology raw data). Informed consent was waived, as the research was observational and used secondary data.

### Advanced haematology data

Whole blood collected in EDTA tubes were analysed using Sysmex XN-1000 and XN-3000 haematology analysers. The XN-3000 is made up of two XN-1000 modules and produces the same haematology parameters. Advanced haematology data was exported from the Information Process Unit (IPU) connected to both a Sysmex XN-1000 at Waitakere hospital and XN-3000 at North shore hospital. This included data from both inpatients and outpatients within Waitemata District Health Board’s (WDHB) catchment. Data was downloaded from the IPU in a comma separated value (CSV) data format, containing both basic and advanced haematology parameters.

The Sysmex XN instrument uses fluorescence flow cytometry, impedance, hydrodynamic focussing, SLS for haemoglobin and is capable of processing up to 100 samples/hour using 88 µL sample volume. Up to 38 clinical parameters and 50 research parameters, or derivatives thereof are produced with up to 23 scattergrams and 4 histograms. The instrument stores up to 100,000 records in a buffer. A glossary of haematology parameter acronyms and explanations are available in a previously published supplemental (22).

### Biochemistry data

Standard biochemical laboratory data from a Siemens Vista 1500 biochemistry analyser, software version is 3.9_11.6-3t/1450, and sent to the laboratory information system (LIS) Éclair and both entered into a national database (Testsafe) and hospital SQL data warehouse. Biochemistry data was extracted from a SQL database using a Python script and matched to either conventional FBC data or advanced haematology data by its closest time point. Biochemistry data at the closest possible time point was matched to either the standard FBC or advanced rawFBC data however as this data was collected as part of clinical care there was significant degree of sparseness/missingness compared to the FBC data. Machine learning models were developed inclusive and exclusive of missing numerics, and with additional clinical data to demonstrate the utility of adding orthogonal multimodal data. NT-proBNP (N-Terminal pro B-type Natriuretic Peptide) was measured using a Siemens Dimension Vista assay and linked by closest time point to data in ECLIPSE. In PROGRAM, NT-proBNP could be linked to a patient encounter but not by blood test date. A ML model, using advanced haematology data, was trained on an abnormal NT-proBNP (≥ 34 pmol/L) to provide corroboration of the features used to predict the coded ICD10 heart failure endpoint.

The ECLIPSE heart failure model was also validated in the PROGRAM test dataset for comparison. The PROGRAM model was validated in an independent dataset inclusive of rawFBC values taken throughout multiple hospitalisations.

### Outcomes

ICD10 primary diagnoses, age, ethnicity and sex were obtained from specific hospital encounter numbers using a SQL query along with mortality data. Congestive heart failure was defined as an ICD10 code within a patient’s entire list of diagnoses. A list of specific ICD10 codes of interest included I509, I500, I130, I110, I132, I42, I25. This also included readmission outcome data. Heart failure with reduced ejection fraction (HFrEF) was defined as an ejection fraction by Simpson biplane < 50% and heart failure with preserved ejection fraction (HFpEF) ≥ 50%. Heart failure machine learning and biological age probability scores (22), were generated for patients in the PROGRAM validation group where there were ≥ two serial blood tests within the same admission encounter. Scores from these two time points (last and first within an admission and discharge) were compared for changes related to disease progression and therapeutic response.

### Statistics and machine learning

Univariate analysis was performed using the student t-test for continuous parametric variables and receiver-operating characteristic curve (ROC) analysis was used to assess performance of diagnostic biomarkers by c-statistic. Z scores were used for proportional values of missingness. All tests were two-tailed and P<0.05 deemed statistically significant, except where Bonferroni correction for multiplicity was applied. Medcalc software version 16.8.4 was used to analyse the data. BigML https://bigml.com/ was used for applying machine learning models, using decision trees, and ensembles, logistic regression and deep neural networks with transparency (https://static.bigml.com/pdf/BigML_Classification_and_Regression.pdf?ver=c306567#page=250). Model development involved splitting data 80:20 into training and test sets. OptiML, an automated BigML optimization process for model selection and parametrization was used to find the best supervised model for sex classification and predicting age using regression. OptiML uses Bayesian parameter optimization and Mote Carlo cross-validation (https://static.bigml.com/pdf/BigML_OptiML.pdf?ver=c306567). An interactive network was generated to compare the rawFBC metadata using a Javascript D3 Force layout and a Pearson correlation matrix. K-nearest neighbour clustering was used for unsupervised machine learning. Uniform Manifold Approximation and Projection (UMAP) was used to visualise clusters in the advanced haematology and biochemistry data from PROGRAM.

### Data availability

The materials, data, code, and associated protocols are available to readers with application to the corresponding author and Waitemata Privacy, Security and Governance (PSGG) group with a limited data sharing agreement. BigML models will be shared without limitations.

## Results

### Machine learning applied to routine laboratory data

ECLIPSE includes data from 75,619 patients who had undergone an echocardiogram. Baseline patient characteristics are in Table 1 and clinical results in Table 2. In patients without heart failure (n = 67,588) the mean (SD) interval between the echocardiogram and the most recent haemoglobin was – 60 (+/-342) days, and between the haemoglobin and creatinine was – 2 (+/-313) days. In patients with heart failure (n = 8,031) the mean (SD) interval between the echocardiogram and the most recent haemoglobin was – 7 (+/- 77) days, and between the haemoglobin and creatinine was + 1 (+/- 72 days). There were 8,031 controls.

**Table 1.** ECLIPSE baseline patient characteristics for heart failure and control groups. Total number of patients with each variable displayed, as well as proportion to total number of patients (%). Age displayed as mean (SD).

|  | Heart failure n = 8031 | Controls n = 67,588 | P Value |
| --- | --- | --- | --- |
| Gender Male (%) | 4,368 (54) | 21,667 (32) | < 0.0001 |
| Age mean (SD) | 76 (13) | 60 (19) | < 0.0001 |
| Hypertension (%) | 5,363 (67) | 11,996 (18) | < 0.0001 |
| Type II diabetes (%) | 2,450 (31) | 5,240 (8) | < 0.0001 |
| Family history of CAD (%) | 93 (1) | 550 (1) | 0.75 |
| Chronic renal failure (%) | 2,617 (33) | 2,420 (4) | < 0.0001 |
| Dyslipidaemia (%) | 1,807 (23) | 4,214 (6) | < 0.0001 |
| ACE inhibitor (%) | 5,568 (69) | 12,600 (19) | < 0.0001 |
| Beta blocker (%) | 6,142 (76) | 14,805 (22) | < 0.0001 |
| Calcium channel blocker (%) | 4,755 (59) | 10,000 (15) | < 0.0001 |
| Statin (%) | 5,469 (68) | 16,323 (24) | < 0.0001 |
| RCA disease > 50% (%) | 1,280 (16) | 3,930 (6) | < 0.0001 |
| LAD disease > 50% (%) | 1,528 (19) | 4,824 (7) | < 0.0001 |
| Cx disease > 50% (%) | 1,201 (15) | 3,475 (5) | < 0.0001 |
| Coronary artery bypass surgery (%) | 82 (1) | 118 (0.1) | < 0.0001 |
| Prior atrial fibrillation (%) | 3,974 (49) | 5,359 (8) | < 0.0001 |
| Future atrial fibrillation (%) | 521 (6) | 855 (1) | < 0.0001 |
| Prior myocardial infarction (%) | 2,637 (33) | 6,164 (9) | < 0.0001 |
| Future myocardial infarction (%) | 339 (4) | 340 (1) | < 0.0001 |
| Mortality (%) | 4,024 (50) | 4,601 (7) | < 0.0001 |

**Table 2.**
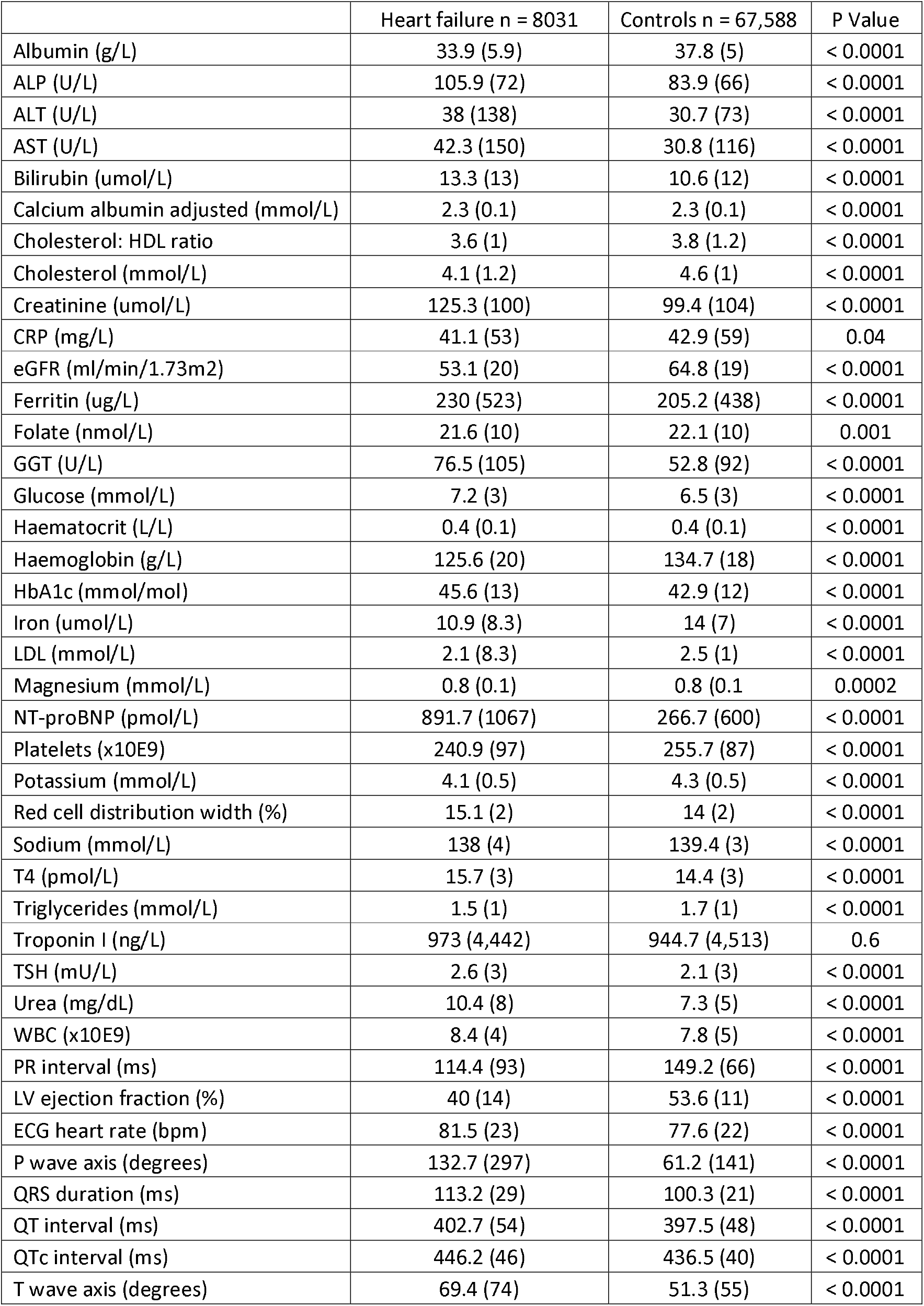
ECLIPSE patient clinical results for both heart failure and control groups. Mean (SD) displayed for each variable and corresponding student t-test P Values.

Only 3,644 (23%) patients of the total 16,062 had an ejection fraction measured by Simpson biplane (EF bp), 2,058 (26%) within the heart failure group. Of those with heart failure and a measured Simpsons ejection fraction the EF bp was < 50% in 1,550 (75%), and < 40% in 1,072 (52%). Despite having no ICD10 code for heart failure within the control group n = 535 (7%) had an EF bp < 50%. These patients were retained within the control group with the presumption that there had been no hospital admission with congestive heart failure.

ECLIPSE included NT-proBNP results for 4,485 patients, 3,913 (56%) of the heart failure group and 572 (7%) of the group without heart failure. Similarly high sensitivity troponin I values were available for 13,081 patients, 7,768 (96%) with heart failure and 5,313 (66%) without heart failure.

12,849 individual patient laboratory results were available for training and 3,213 for testing ML models. A deep learning model applied to just age + conventional bloods including missing numerics had the highest accuracy predicting heart failure (Table 3), with highest ranked features being haematocrit (32%), age (24%), red cell distribution width (RDW) (18%), T4 thyroxine (4%) and troponin I (4%). However incremental gains were demonstrated with the addition of further demographics (ethnicity), orthogonal information (conventional ECG parameters: PR interval, QRS duration and T wave axis) and longer training time. Model performance reduced substantially with the exclusion of missing numerics, suggesting a possible role of bias. Three (14%) of the 22 laboratory variables had statistically greater proportions of missingness in the control group (red cell distribution width, cholesterol HDL ratio and HbA1c) compared to those with heart failure. Various machine learning methods e.g. decision tree ensemble, deep learning and logistic regression provided the most accurate model, based on phi score and this was not limited to just deep learning. Even in the absence of key variables, e.g. RDW, reasonable predictions were still possible.

**Table 3.** Accuracy of ECLIPSE machine learning models

| Machine learning models | AUC | SE <sup>a</sup> | 95% CI <sup>b</sup> |
| --- | --- | --- | --- |
| (1) Conventional bloods + ethnicity + ECG + long train time | 0.929 | 0.00436 | 0.919 to 0.938 |
| (2) Conventional bloods + ethnicity + ECG + short train time | 0.925 | 0.00451 | 0.915 to 0.934 |
| (3) Conventional bloods, including missing numerics | 0.921 | 0.00466 | 0.911 to 0.930 |
| (4) Conventional bloods + ethnicity, excluding missing numerics | 0.870 | 0.00612 | 0.858 to 0.882 |
| (5) Conventional bloods, excluding missing numerics | 0.851 | 0.00662 | 0.838 to 0.863 |

<sup>a</sup> DeLong et al., 1988
<sup>b</sup> Binomial exact

Conventional blood ML model accuracy was compared with NT-proBNP with comparable results, though the ML model including missing numerics exceeded NT-proBNP’s accuracy in identifying heart failure, AUROC 0.87 versus 0.81, 95% CI 0.004 to 0.0974 P = 0.03 (Figure 3).

**Figure 3.**
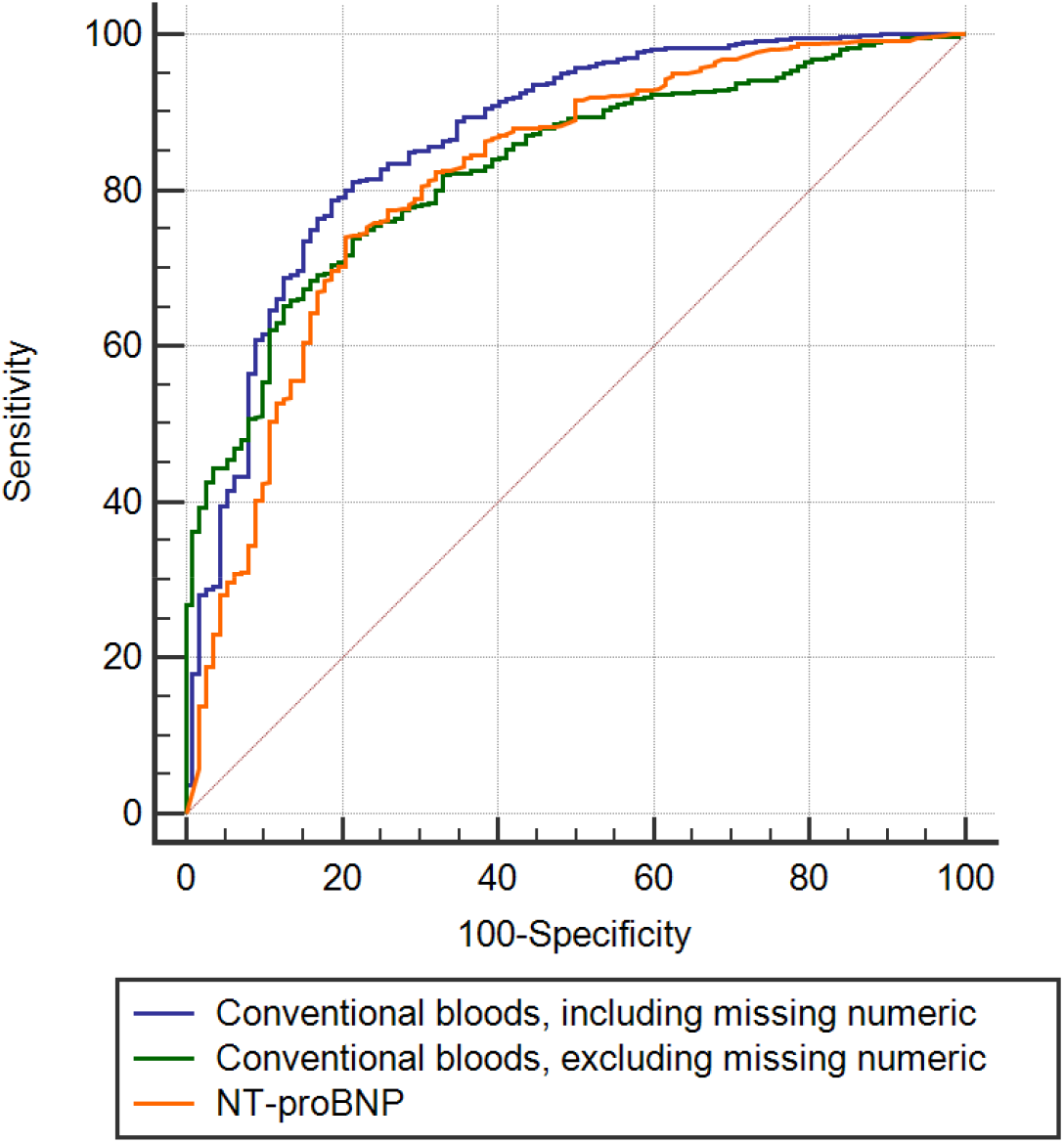
Receiver operator curves comparing ECLIPSE models with NT-proBNP

678 (21%) of patients in the test set had EF bp measured, 294 with HfpEF and 383 with HfrEF. Applying both the conventional bloods ML models, including and excluding missing numerics, demonstrated similar accuracy for identifying both HfrEF AUROC 0.88 to 0.79 (95% CI 0.05 to 0.13, P < 0.0001) and HfpEF AUROC 0.91 to 0.83 (95% CI 0.04 to 0.11, P = 0.0001). Including NT-proBNP in the HfpEF ML model increased the AUROC to 0.93, with sensitivity 86% and specificity 85%, 95% CI 0.89 to 0.95, P < 0.0001. Using the entire ECLIPSE dataset, both past and future HfpEF (n = 416) compared to controls (n = 6,800 controls; EF bp ≥ 50%, no heart failure) was predicted with AUROC 0.997, 95% CI 0.992 to 0.999, P < 0.0001. Highest ranked features were two chamber stroke volume (SV) (17%), HbA1c (5%), left ventricular end diastolic dimension indexed to height (4%), Teicholz SV (4%), potassium (4%). Other predictive models with similar accuracy included ejection fraction, QTc, and other variables. Echocardiography variables dominated the highest ranked features for HfpEF prediction in most models.

k-nearest neighbour unsupervised machine learning was applied to conventional bloods, ECG and echocardiography parameters in 400 patients with HfpEF. Three phenoclusters (Figure 4 and Table 4) were identified with variable longitudinal mortality outcomes, replicating work by Shah et al and others (16, 17).

**Figure 4.**
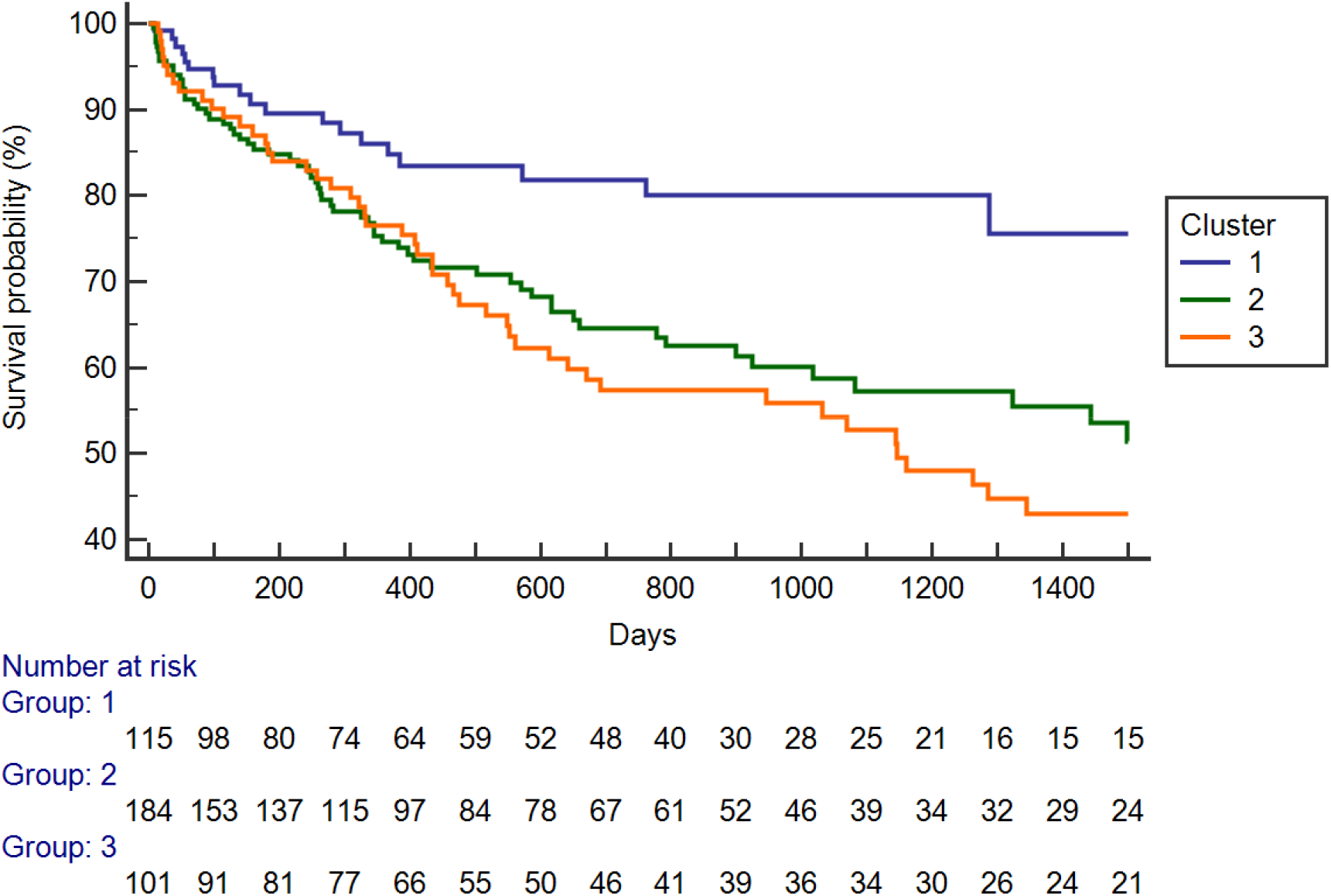
Kaplan Meier plot of HfpEF clusters

**Table 4.** Cluster phenotype patterns

|  |  |  |  |
| --- | --- | --- | --- |
| Cluster 1 | Larger heart | Higher SV | Obese, less HTN, dyslipid, younger, lowest NT-proBNP |
| Cluster 2 | Smaller heart | Lower SV | Long PR interval |
| Cluster 3 | Diastolic dysfunction | Largest LA | Older, renal failure, Low rate T2DM, longest p wave |

**Table 4.** Accuracy of PROGRAM machine learning models

| Variable | AUC | SE <sup>a</sup> | 95% CI <sup>b</sup> |
| --- | --- | --- | --- |
| (1) Advanced haematology + biochemistry | 0.925 | 0.0234 | 0.868 to 0.963 |
| (2) Advanced haematology | 0.913 | 0.0233 | 0.853 to 0.954 |
| (3) Conventional bloods, including missing numerics | 0.832 | 0.0366 | 0.759 to 0.889 |

<sup>a</sup> DeLong et al., 1988
<sup>b</sup> Binomial exact

### Machine learning applied to advanced haematology and biochemistry data

314 heart failure patients and 384 controls were used for training and testing machine learning models using age + advanced haematology (rawFBC) and conventional biochemistry data. Incremental gains were demonstrated with both the use of advanced haematology data and biochemistry data Table 4, compared to the ECLIPSE model (95% CI 0.03 to 0.15, P = 0.002) Figure 5. Although there appeared to be some gain adding biochemistry data to advanced haematology this comparison was not statistically significant, possibly due to underpowering. Highest ranked features in the ML model including advanced haematology data were age (18%), RDW-SD(fL) (10%), RDW-CV(%) (7%), MacroR(%) (5%) and NEUT#(10^9/L) (4%) demonstrating similar features to ECLIPSE, e.g. RDW, and pathophysiological features of heart failure consistent with prior literature. Highest ranked features in the combined advanced haematology and biochemistry model included creatinine (9%) but highest ranked features were still dominated by haematology variables.

**Figure 5.**
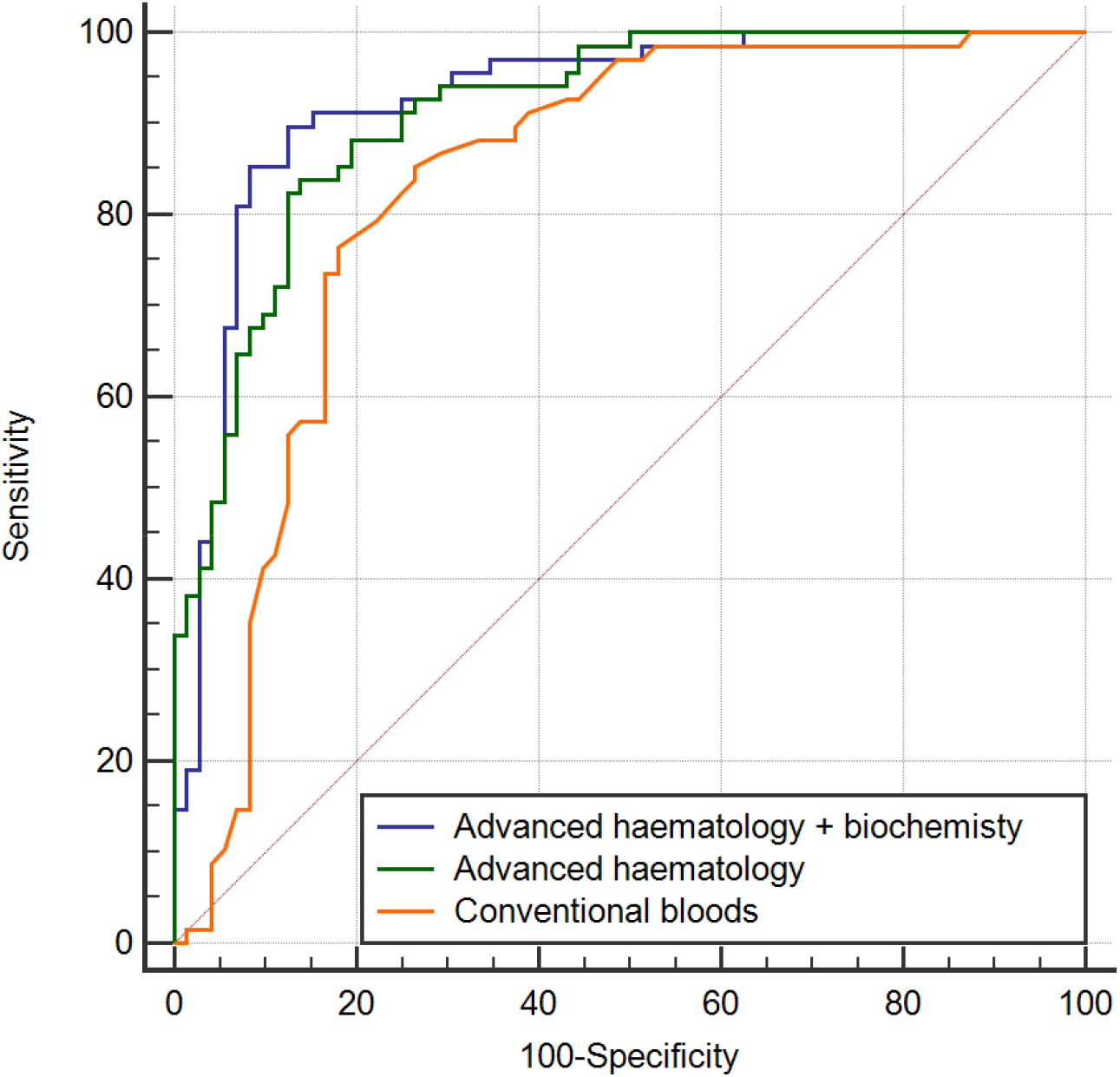
Receiver operator curves for PROGRAM and validation of ECLIPSE n = 140

In the PROGRAM validation set there were 69,492 FBCs, 2.3% of which had an ICD10 primary diagnosis of heart failure. Heart failure was predicted with a sensitivity 75%, specificity 76% (Figure 5), positive predictive value 7%, negative predictive value 99.2% (AUROC 0.83, 95% CI 0.83 to 0.84, P < 0.001) in validation (Figure 6).

**Figure 6.**
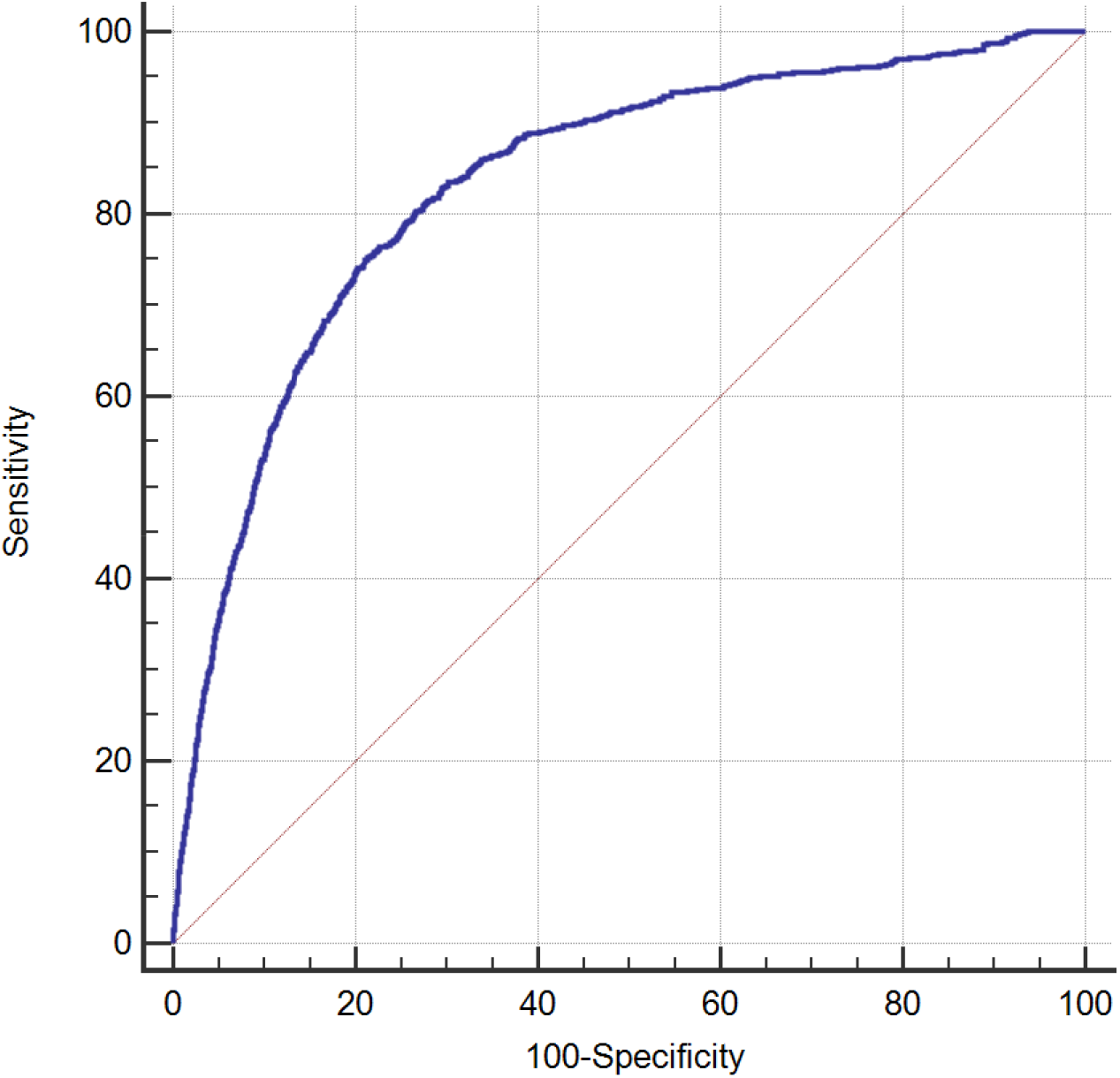
Validation of PROGRAM, machine learning heart failure model

1,028 rawFBC results had an associated NT-proBNP ≥ 34 pmol/L during the same hospital encounter 452 (44%). A decision tree ensemble predicted an NT-proBNP ≥ 34 pmol/L with AUROC 0.97 (95% CI 0.93 to 0.99, P < 0.0001). Red blood cell agglutination (9%), haemoglobin (g/L) (8%), haematocrit (7%), RDW-CV(%) (6%), haemoglobin defect (4%) were the highest ranked features consistent with models predicting ICD10 codes for heart failure.

### Network analysis

Visualisations of the advanced haematology and biochemistry data from PROGRAM is available as an interactive network here https://projects.interacta.io/theranostics-covid/.

UMAP clustering of combined rawFBC and biochemistry data showed distinct separation of patients with heart failure with controls (Figure 7).

**Figure 7.**
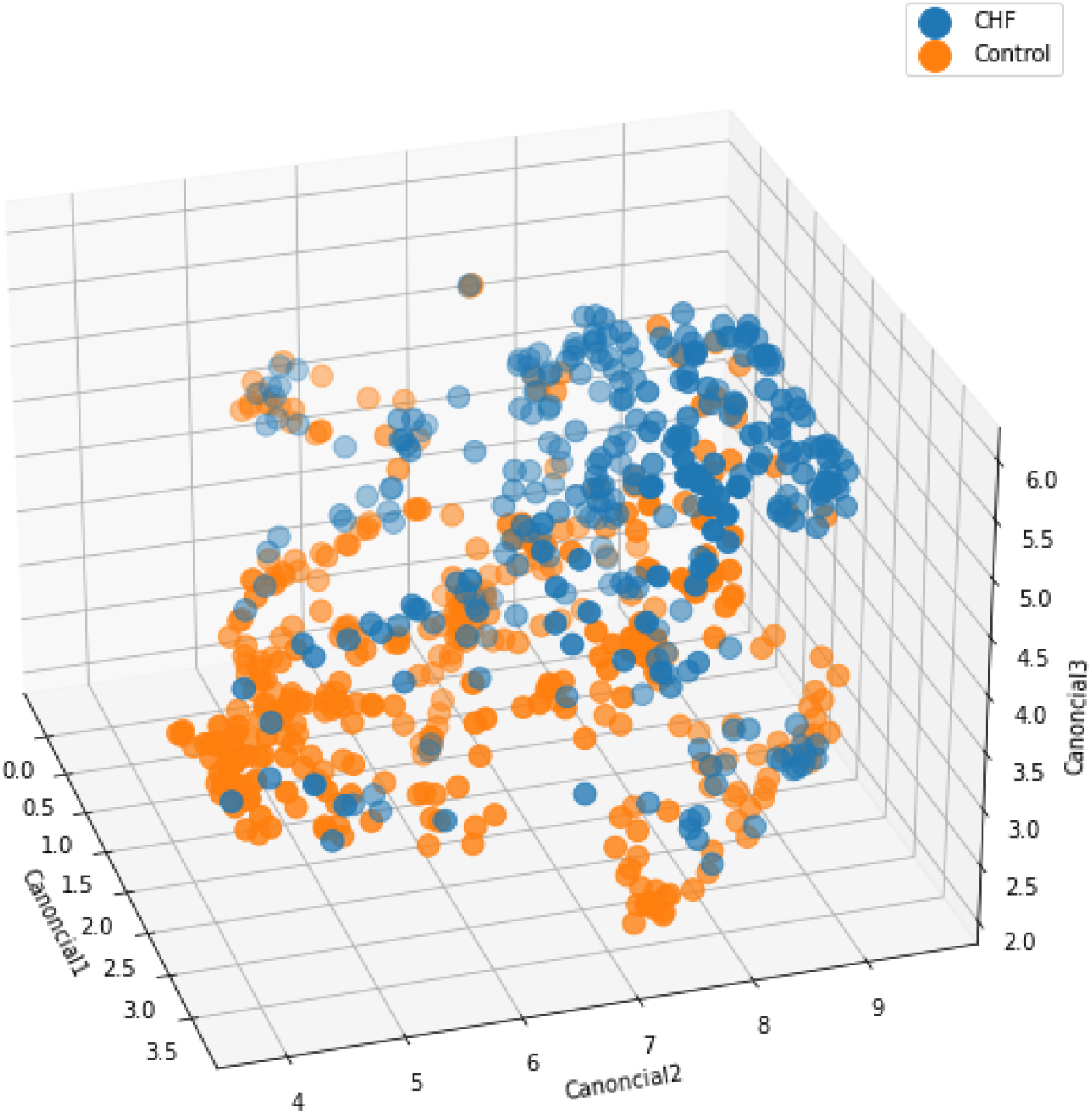
UMAP clustering of PROGRAM data

### Global and Individual patient responses to treatment

We calculated a heart failure disease probability score and biological age score for each blood result to compare an admission and discharge result. Although there was considerable individual heterogeneity in haematology heart failure patterns over time, the mean global difference between these two time points was not statistically significant (Figure 8a and 8b). The individual response to dapagliflozin in a single patient was demonstrated using only two haematology parameters (Figure 9).

**Figure 8a and 8b.**
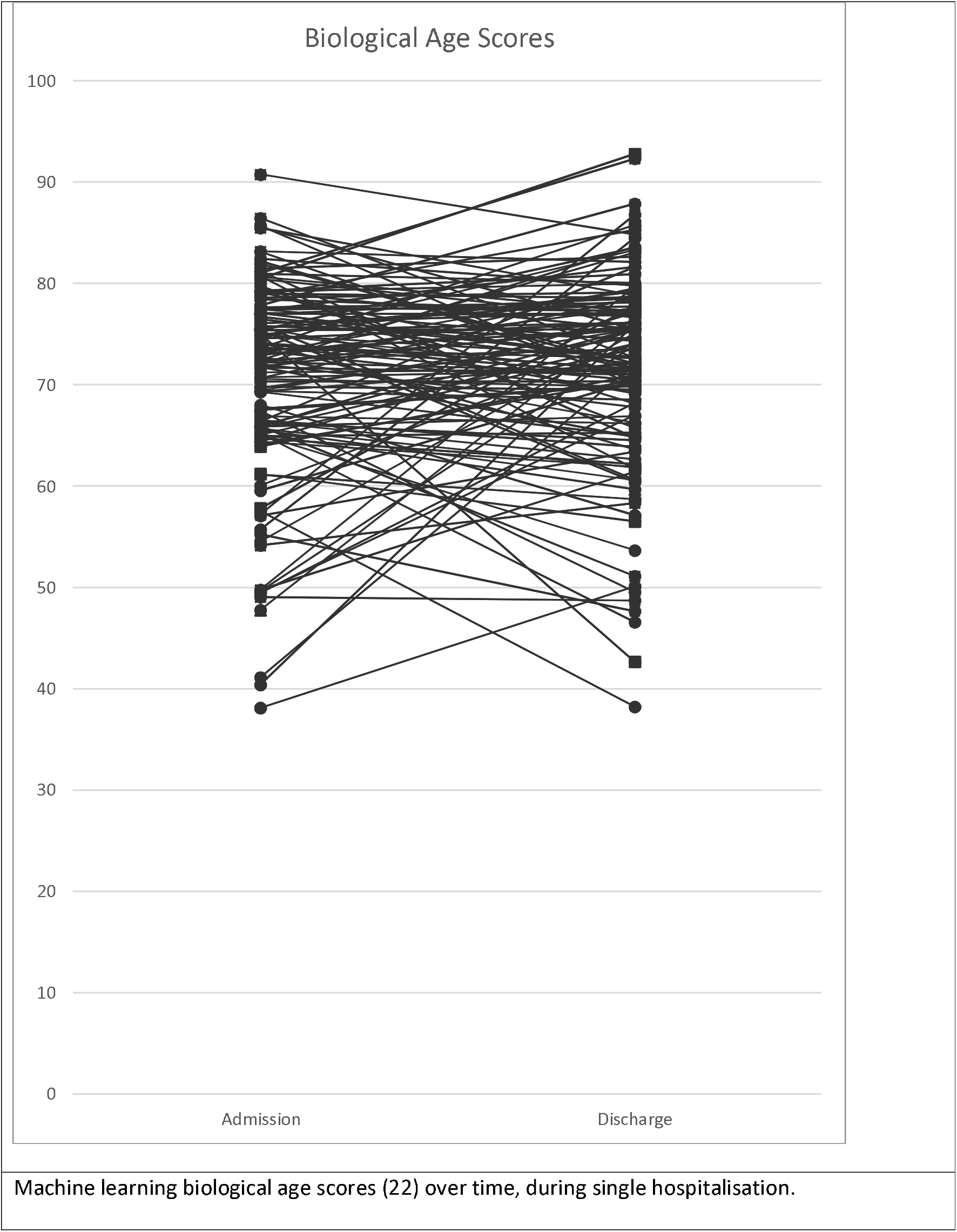
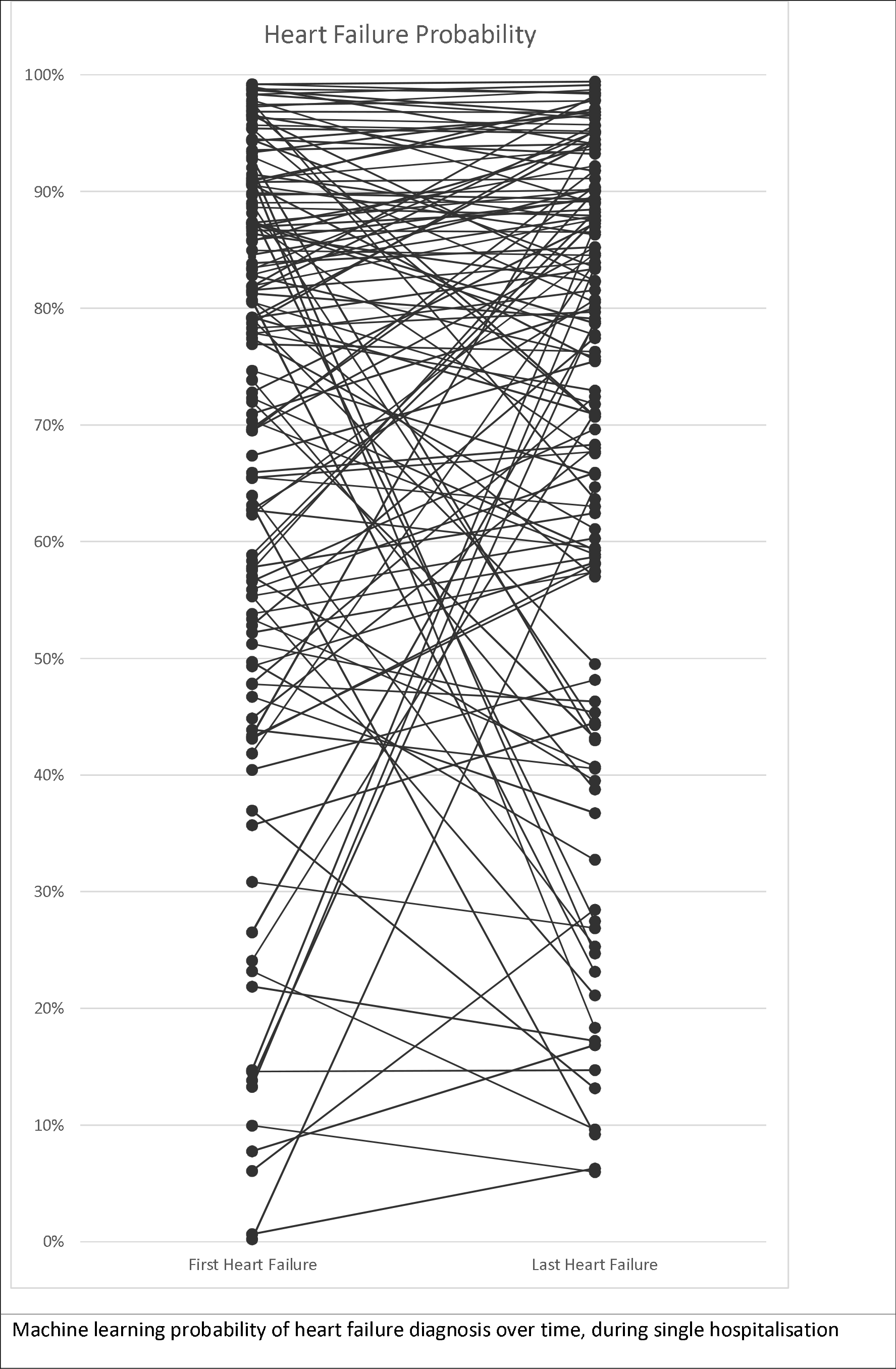
Machine learning scores for biological age and heart failure over time for individual patients, during single hospitalisation

**Figure 9.**
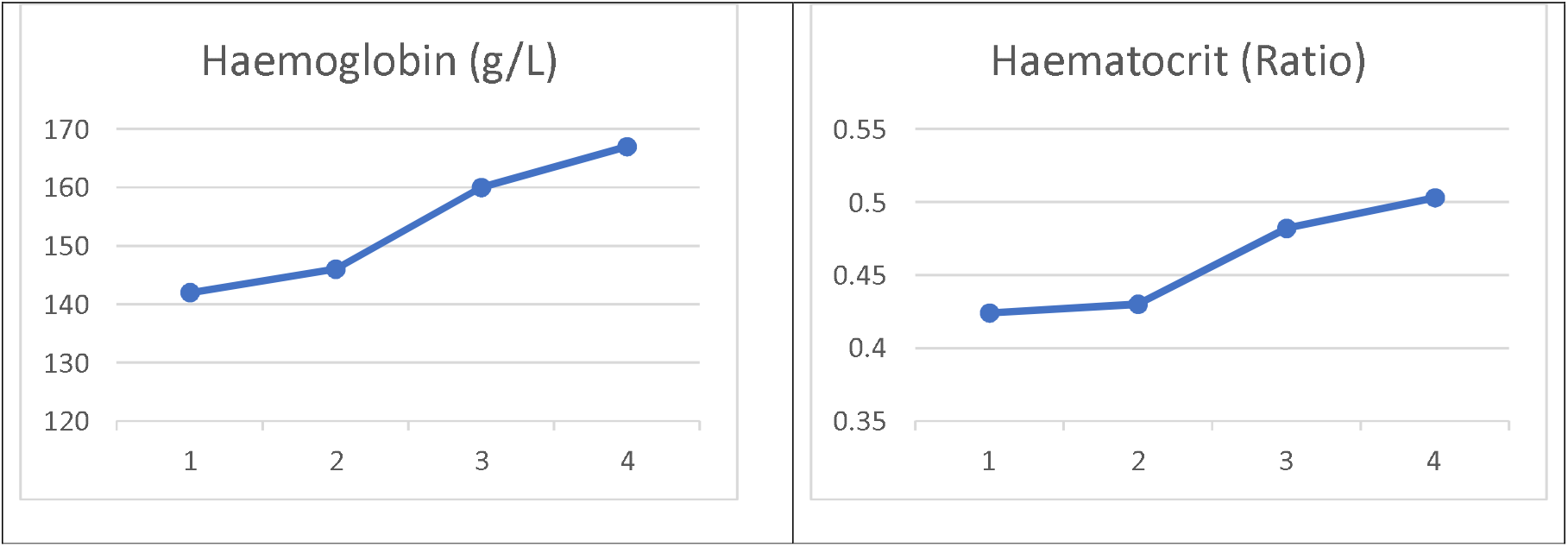
Individual response to dapagliflozin, started at timepoint 2.

## Discussion

In these studies (ECLIPSE and PROGAM) we have demonstrated it is possible to use machine learning to predict the presence of heart failure from both conventional bloods and advanced haematology data. We used an automated Cloud-based machine learning, software as a service platform, which required no coding experience. The machine learning methods varied between decision tree ensemble, logistic regression and deep learning however, no particular method exceeded others in predictive accuracy. What was apparent is that by increasing compute time and the addition of orthogonal data (e.g. ethnicity and ECG data) incrementally improved model predictions, though with diminishing returns. Although the machine learning models were transparent and explainable, with ranked identifiable features, these did not aid in making causal inferences with each predictive model. For instance, although ethnicity provided incremental value in predictions it cannot be assumed that specific at risk ethnicities carry a higher risk due to genetic variants (such as titin truncations, which increase the risk of heart failure) or if socioeconomic confounding is having an influence (23). Another concern with including non-biologically causal predictive features is the risk of dataset shift and bias (24).

A note of caution must be made in placing excessive confidence in ECLIPSE predictions combining haematology and biochemistry, for the reason that whilst some bloods are ordered almost universally, such as an FBC, others e.g. iron studies are only ordered on the basis of a preconceived diagnostic notion or concurrent condition. Therefore, the presence or absence of data in itself represents a source of self-fulfilling bias (25, 26). Furthermore, health care data collected in a real-world setting is plagued with not only missing data but also time variance, meaning training data may not always be collected prior to the confirmation of a diagnosis. These encoded hidden biases in the data could easily have skewed test results into demonstrating predictions of much higher accuracy, as possibly demonstrated with the removal of missing numerics from the predictive model. The control group from which the ECLIPSE data was randomly selected had a far higher frequency of missing data, possibly due to being ambulatory patients rather than inpatients. However even after accounting for this bias, by excluding missing numerics, the model still performed well on an independent inpatient dataset within PROGRAM and when compared with NT-proBNP within a subset of the ECLIPSE patients.

In PROGRAM, machine learning models discriminated heart failure from both rawFBC and rawFBC plus biochemistry with similar accuracy to the ECLIPSE models. The results were underpowered to demonstrate improved accuracy with the addition of biochemistry data, however the results from just the rawFBC are interesting for a number of reasons. Firstly, the ability to discriminate heart failure with an approximate AUROC 0.83 has now been validated internally twice (22) and independently (21). Secondly, since the data exported from the Sysmex analyser is complete, for each sample, missing values and bias is minimised.

Thirdly, Sysmex haematology analysers are in widespread use, in over 85% of clinical laboratories, and produce highly consistent results independent of geographic location etc. Others have indicated that machine learning predictions, based on haematology data, could be used for adjudicating endpoints for more encompassing machine learning applications, or by institutions or payers to monitor population level data (21). We would concur with that, with the addition that such technology could be useful at both an individual level in predicting risk (27), particularly in resource constrained settings but also a population scale for the surveillance and monitoring of not only noncommunicable cardiovascular disease but also infectious diseases and pandemics (22).

Two factors increase the confidence in the validity and clinical utility of both the results from PROGRAM and ECLIPSE. Firstly, the data in PROGRAM was collected from a single snapshot in time at the first point of contact in an admission, unlike ECLIPSE. Further work will be required to demonstrate incremental diagnostic gains from the use of machine learning predictions applied to haematology data, however since heart failure, particularly HFpEF, is often undifferentiated at presentation the likelihood of this adding value either in the emergency department or primary care is high. Secondly, when interrogating the highest ranked features of the predictive models in both ECLIPSE and PROGRAM some commonalities are clearly seen. Markers of erythropoiesis, such as red cell distribution width (RDW), haemoglobin concentration and haematocrit, clearly dominate the statistical models, explaining why haematology data on its own is capable of making fairly accurate predictions.

Whilst anaemia in heart failure has been well documented, its causes are multifactorial in part due to co-existent renal disease, iron storage and transport and failure of erythropoietin production (28). Similarly altered haematocrit, commonly seen in heart failure and presumed due to haemodilution, has been identified as both a predictor (29) and risk factor in heart failure (30). Paradoxically in a population setting a higher haematocrit is associated with an increased risk of heart failure (29). This may be related to causal ABO genetic variants, themselves associated with lipid traits identified through genome wide association studies (GWAS) of haematological quantitative traits (31-33). Therapeutic targeting of erythropoiesis in heart failure has had mixed results, though not for iron replacement therapy (28, 34). Recently SGLT2 inhibitors have been shown to have significant benefits in both HFrEF and HFpEF (35), with effects predicted by erythropoietic responses and their ability to alleviate cellular stress (36, 37). These effects seem to operate at the level of aging related pathways such as SIRT1 and AMPK (38, 39) possibly indicating a role beyond heart failure and diabetes. Abnormal red cell distribution width (RDW), a marker of loss of control of homeostasis otherwise known as anisocytosis, was identified here as a key feature in heart failure prediction models. RDW has been well described as a predictor of heart failure outcomes (40) and a GWAS has similarly demonstrated its ability to predict a range of negative health outcomes is similarly influenced by fundamental pathways of aging (41). We and others have previously demonstrated RDW’s role in the prediction of biological age using ML applied to blood results (22, 42). Although we were not able to show serial changes in either the prediction scores for heart failure or biological age at a population level, this does not rule out the possibility of these being useful at an individual level particularly in monitoring therapeutic responses to heart failure therapies, such as SGLT2 inhibitors (Figures 8 and 9). We have previously shown an ability to monitor LV systolic dysfunction over time, using personalised trajectories, using ML applied to ECG (2), and integrated this with multiomics, to provide a more holistic overview of HFrEF (3).

With the expanded global metadata also available in ECLIPSE we have shown added value of incorporating heterogenous clinical data into machine learning models to predict certain diagnoses, such as HFpEF with higher accuracy. Although ML applied to conventional bloods predicted both HFrEF and HFpEF with reasonable accuracy, blanket application of such a tool at population scale would result in a larger number of false positives, more so than true positives due to the low prevalence of heart failure (2.3%) in hospital blood results, shown in our validation set of 69,492 FBCs. Targeting this technology in a sequential manner, possibly starting with a standard clinical history but then adding in more costly investigations such as NT-proBNP, electrocardiography and echocardiography could result in better use of resources, as recently shown by a Mayo group investigating the use of deep learning applied to ECG (43, 44). A predictive model for HFpEF in ECLIPSE was dominated by echocardiography variables, notably stroke volume and others indicating that echocardiography is an essential component to the identification of HFpEF. Global clinical metadata was also useful in validating prior work done in subtyping HFpEF into various phenogroups, using unsupervised machine learning (16, 17). Unsupervised machine learning holds significant promise in extracting latent space from high dimensional clinical data to reclassify the taxonomy of disease. This may have particular utility in circumstances where clinician driven diagnostic coding does not capture the full nuance or longitudinal spectrum of a complex condition, such as HFpEF. With the removal of key features from machine learning models, reasonable predictions were still possible using global clinical metadata suggesting a self-healing, almost holographic quality of broad clinical data.

## Conclusion

We have demonstrated in this study an ability to discriminate heart failure, both HFpEF and HFrEF using machine learning applied to both routine bloods as well as advanced haematology data, extracted from a commonly used flow cytometer. The predictions obtained from the haematology flow cytometer have been validated independently and due to their underlying predictive features might have value in predicting and monitoring responses to heart failure therapies in near real-time. The use of machine learning applied to abundant, low-cost laboratory data may have particular value in low to middle income countries where access to high level, expensive diagnostic testing is limited.

## Data Availability

Anonymised data from this study is available on request from the corresponding author, pending approval from local research boards at their respective institutions
Freely available data is provided access via URLs within the manuscript

## Limitations

As with most machine learning studies the results from this project may not be transferable into other settings, due to differences in regional practices, disease presentation and prevalence etc. However transferable machine learning models are preferably generated from data which comes from a similar source, is complete and is resilient to clinician biases. Haematology and to a lesser degree biochemistry data fulfils that requirement. Over-fitting is always a possibility when using data from a single source and preferably we would have applied our ML models to external data to validate. The use of matched cases and controls over-estimates the accuracy of predictive models when using AUROC (45), however we also validated our models in a real world dataset to demonstrate what implementation might look like. Clinical utility cannot be assumed until predictive models of heart failure are able to identify missed cases of heart failure or alert to the diagnosis of heart failure earlier, whereby patient outcomes can be improved. Although algorithms applied to existing clinical data may be low-cost to implement the downstream human and societal cost implications of false positives and negatives needs to be studied during an implementation program.

